# Finding hotspots: development of an adaptive spatial sampling approach

**DOI:** 10.1101/2020.01.10.20016964

**Authors:** Ricardo Andrade-Pacheco, Francois Rerolle, Jean Lemoine, Leda Hernandez, Meïté Aboulaye, Lazarus Juziwelo, Aurelien Bibaut, Mark van der Laan, Benjamin Arnold, Hugh Sturrock

## Abstract

The identification of disease hotspots is an increasingly important public health problem. While geospatial modeling offers an opportunity to predict the locations of hotspots using suitable environmental and climatological data, little attention has been paid to optimizing the design of surveys used to inform such models. Here we introduce an adaptive sampling scheme optimized to identify hotspot locations where prevalence exceeds a relevant threshold. Our approach incorporates ideas from Bayesian optimization theory to adaptively select sample batches. We present an experimental simulation study based on survey data of schistosomiasis and lymphatic filariasis across four countries. Results across all scenarios explored show that adaptive sampling produces superior results and suggest that similar performance to random sampling can be achieved with a fraction of the sample size.

## Introduction

Recent years have seen considerable success towards control and elimination of a range of globally important infectious diseases. For many of these diseases, decisions relating to interventions are made across administrative units. For example, decisions about where to conduct mass drug administration campaigns for neglected tropical diseases (NTDs) are made at an implementation unit (IU), typically the district or sub-district level [1]. A similar approach is typically taken in the control and elimination of malaria, where entire districts or sub-districts may receive insecticide treated nets or indoor residual spraying where others do not.

For NTDs, decisions relating to MDA are based on infection prevalence estimates at the IU level obtained from cross sectional surveys. Where IU level prevalence exceeds a threshold, the entire IU is treated [1]. Where prevalence does not exceed this threshold, the IU does not qualify for MDA and no individuals in that area are treated. For example, for schistosomiasis, current guidelines recommend that MDA is conducted in areas where prevalence is greater than 10%, whereas for soil-transmitted helminths, this threshold is 20% [1].

While operationally straightforward, this approach ignores any within IU heterogeneity. In many instances, districts with prevalence below the threshold that triggers intervention contain a number of villages with active transmission [2]. Modeling and intuition therefore suggest that as disease transmission declines, moving away from decision making at coarse scales towards a more targeted approach is more cost-effective [3]. Such targeting is predicated on sufficiently accurate information on the location of sites with an infection prevalence above a policy relevant threshold, from hereon referred to as *hotspots*.

Missing hotspots could cause setbacks for elimination efforts. Hence, various approaches to identify them have been proposed. Variations of contact tracing, whereby testing is targeted at families and neighbours of individuals found positive during surveys or routine surveillance, have been explored for a number of diseases including schistosomiasis [4], lymphatic filariasis [5] and malaria [6, 7]. Such approaches can, however, be expensive and can still fail to identify hotspots if positive individuals from those communities are not identified by the initial surveys.

An alternative approach is to use less costly survey methods to sample a higher proportion of locations than would otherwise be possible. Techniques such as lot quality assurance sampling, a method designed to minimize sampling effort in order to categorize outcomes over a given population, is one such approach and has been used to identify hotspot communities for schistosomiasis [8, 9]. Similarly, school-based questionnaires relating to blood in urine and eye worm occurrence, have been used to map urinary schistosomiasis [10, 11, 12] and loa loa [13, 14] respectively. These methods are inherently noisy as they only allow measurement of proxies of infection and can suffer from issues of recall.

Another approach to mapping hotspots, which negates the need to sample a large fraction of the population, is using geospatial modeling. Climatological, environmental and ecological layers can help predict the spatial distribution of many infectious diseases. Furthermore, above and beyond patterns that can be explained by these layers alone, disease outcomes often display some spatial structure, with neighbouring values being correlated due to shared characteristics and transmission. This spatial structure means that information from one site provides information about neighbouring sites. Over the past decade, the ability to predict pathogen infection prevalence across entire regions based on survey data and relationships using geospatial modeling has improved considerably [15, 16, 17]. These advances in geospatial modeling have opened the door to more targeted approaches, potentially allowing decisions about treatment to be made with higher precision and granularity.

Despite these advances, surprisingly little attention has been paid to optimizing the survey design for risk mapping efforts. Evidence from other fields has shown that random sampling is suboptimal for spatial prediction [18, 19, 20, 21]. For lymphatic filariasis, a grid sampling approach has been proposed as a mechanism to allow for more efficient spatial interpolation [22, 23]. Diggle and Lophaven (2006) propose the use of grid sampling supplemented with clusters of close pairs of points to allow for better estimation of the variogram used for Kriging [24]. Simulation studies also suggest that this design provides a more cost-effective approach to mapping schistosomiasis [3].

More recently, Chipeta et al (2016) and Kabaghe et al (2017) proposed spatially adaptive designs that leverage information from prior data to inform the locations of future sampling sites to minimize prediction error [25, 26]. Using malaria as an example, results from simulations and field studies show that adaptive spatial designs can be used to produce more precise predictions of infection prevalence using geostatistical modeling [25].

Building on the adaptive spatial sampling approach, we incorporate ideas from Bayesian optimization theory to propose an adaptive spatial sampling approach optimized to identify hotspot communities [27, 28].

## Methods

### Spatial Model

To predict the probability that a given site (e.g. a village or other type of settlement) is a hotspot or not, and to guide adaptive sampling schemes, requires fitting a spatial model to observed data. As a reminder, here a hotspot is defined as a location where infection prevalence is greater than a defined threshold. We assume that an initial representative population sample exists to allow a model to be fit. If this is not the case, a randomly sampled set of measurements would be one option, although there may be superior approaches, particularly if data relating to the expected spatial structure or covariate values at candidate survey sites exist [24, 29, 30].

There are a range of different modeling approaches available to predict prevalence at unsurveyed sites. Here, we use a combination of machine learning and model-based geostatistics [15, 31].

Let ℬ be a region (e.g. a country) where we are interested in determining if a set of sites are hotspots or not. As mentioned above, it is assumed that an initial dataset from which we can estimate the overall prevalence exists. Say we have the dataset 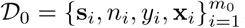, where **s**_*i*_ are the GPS coordinates that describe the location of a site of interest, *n*_*i*_ is the number of people tested in such site, *y*_*i*_ are the number of positive cases out of *n*_*i*_ and **x**_*i*_ are other features associated to the site, like elevation, distance to water bodies or average temperature; *m*_0_ is the total number of observations. Given these data we can model the prevalence in ℬ as a spatially continuous process given by

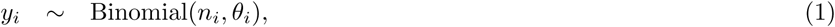

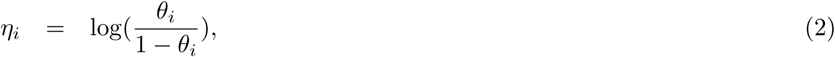

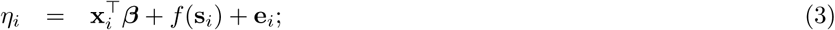

where ***β*** are a set of real parameters and *f* is a spatially correlated random effect using a Matérn correlation function (see Appendix equation 8) and *e*_*i*_ is a residual independent error term.

Instead of including linear covariate effects, we first fit a random forest model using 20-fold cross validation using all the covariates, excluding latitude and longitude. For each observation, we then have a cross-validated prevalence prediction (from hereon termed out-of-sample predictions). Additionally, we fit a random forest using all observations and use this model to predict to all observation and prediction points (from hereon termed *in-sample predictions*). Out-of-sample predictions from the random forest are then included as a single covariate in the geostatistical model (equation 3).

When making predictions, in-sample predicted prevalence values from the random forest using all observations were used as the covariate at each prediction point. While this model allows us to predict prevalence across the continuous region ℬ, in this case we are only interested in predictions at the location of human settlements. Here, we denote these discrete locations as 𝒮 ⊂ ℬ.

In addition to obtaining estimates of predicted prevalence, the model described above allows us to estimate the *exceedance probabilities*, i.e. the probability that prevalence *θ*_*i*_ at location **s**_*i*_ is above a given threshold *ϑ*.

### Adaptive Sampling

#### Exploitation

The goal we seek when using adaptive sampling or adaptive design is to leverage the information available and select the *optimal* sampling locations to improve our statistical inference [27, 28]. The criteria to define what is optimal depends on what quantity is to be estimated. Hence, it is first necessary to define an objective or utility function, i.e. the measure by which we evaluate the performance of any given design. For situations where the goal is to produce as precise predictions as possible over the study region, measures such as average prediction variance is a sensible option [25]. If, however, the goal is to find hotspots, we are less interested in the precision of our estimates and should be focused on minimizing hotspot classification error from our model. Put another way, we wish to increase our confidence that the prevalence at any given location is above or below the predefined threshold. A measure that fits naturally into this framework is *Shannon entropy*. Shannon entropy measures the uncertainty of a random variable based on its probability distribution [32]. Let *ϑ* be the relevant threshold. Given the model described in Eq 1, for every **s**_*i*_ ∈ 𝒮 we can estimate its probability of being a hotspot *p*(*θ*_*i*_ *> ϑ*|𝒟_0_). Then the entropy value at such location regarding it being a hotspot or not is defined as

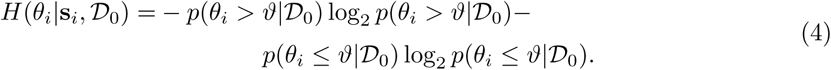

Locations with exceedance probabilities of 0.5 (i.e. 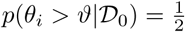) are the most uncertain and have an entropy value of one. On the contrary, the more certainty in the event (i.e. exceedance probabilities close to 0 or 1), the entropy gets closer to 0. By targeting high entropy values, sampling is focused on those sites with highest classification (hotspot or not) uncertainty.

#### Exploration

Giving preference to locations with higher uncertainty is intuitively more efficient than a uniform random selection, but choosing the design based only on entropy values (Eq. 4) may not be efficient because prevalence is usually a spatially correlated process. For example, see Figure 1 panel A, where we show a simulated field of uncertainty where values are spatially correlated. Since locations with high uncertainty can be expected to be clustered together, by defining a batch of sample points based only on *H*(*θ*_*i*_|**s**_*i*_, 𝒟_0_) we may end up selecting locations that are very close to each other. However, such an approach leads to redundancy, as taking a measurement at one location also provides information about neighboring locations due to the spatial correlation present. In Figure 1 panel B, we choose the 10 locations (red dots) with highest uncertainty values from a grid of 15 *×* 15 potential locations (white dots). The Figure demonstrates how this greedy approach can result in poor coverage of the field.

**Figure 1.**
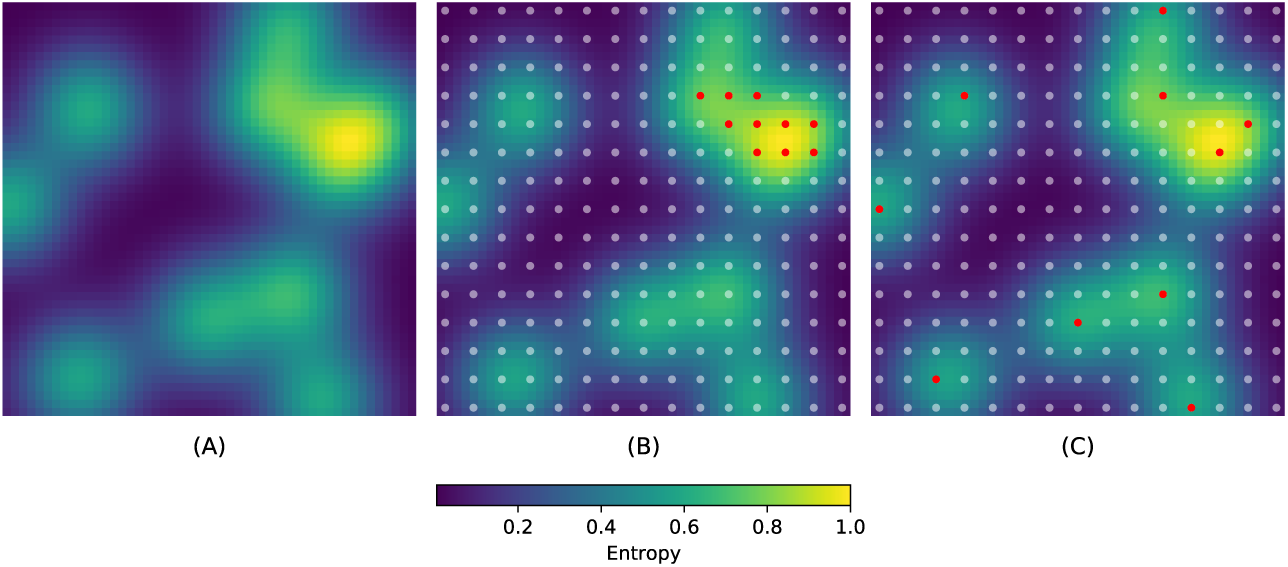
Exploration-exploitation trade-off. A: Spatially correlated uncertainty. B: Batch selected (red dots) by using the greedy approach of targeting the highest values of uncertainty. C: Batch of locations selected (red dots) using the acquisition function described in Eq. 7.

It would be preferable to sample high entropy points, while ensuring a good spread of points across the study area to avoid redundancy. This allows a balance between exploitation (i.e. targeting high values of *H*(*θ*_*i*_|**s**_*i*_, 𝒟_0_)) and exploration (i.e. spread batch locations in ℬ) [33]. If in Eq. 3 we assume that *f* is a multivariate Gaussian with spatial covariance **K**(**s**_*i*_, **s**_*j*_), then the average amount of information contained in a batch of locations 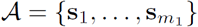 is given by the joint *differential entropy*

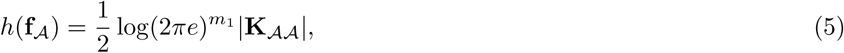

where 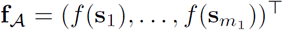 and **K**_𝒜 𝒜_ = [**K**(**s**_*i*_, **s**_*j*_)].

The differential entropy is the continuous case of the Shannon entropy introduced before [32]. A low value of *h*(**f**_𝒜_) implies that the random variable **f**_𝒜_ is confined to a small volume, whereas a large value of the differential entropy implies a that the variable is widely dispersed. Given a batch size, by choosing the elements in it that maximize the differential entropy, we would be maximizing the average information content of the batch with respect to the random field *f*. Finding the batch with highest information content is a problem of combinatorial complexity. However an exact solution is not needed [34]. A approximate solution can be found through a sequential approach that at step *t* selects the new element of the batch according to

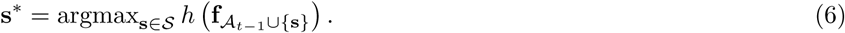

#### Trade-off

Once we have a utility function and a rule for exploration, we only need to define a trade-off strategy between exploration and exploitation that helps us select a batch of new survey locations. In Bayesian optimization, this strategy is defined by the *acquisition function* [35, 36]. Notice, however, that our setting is simpler than the usual setting for Bayesian optimization, where evaluating the utility function is considered to be expensive and the exploration sites could be infinite. In this application we assume a finite set of potential survey locations, as they represent villages or some type of human settlements. Also, in all of these locations we have a measurement of our utility function through the posterior distribution of *θ*.

As trade-off strategy we define the step-wise algorithm that combines Eq. 4 and Eq. 6, so that at step *t* the new element in the batch is chosen according to

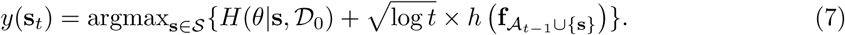

In the expression above we are explicitly defining *y* as a function of **s** to emphasize that we are interested in selecting survey locations. By using this acquisition function we induce batch locations to be spatially scattered and therefore achieve a better exploration. In Figure 1 panel C, we show a batch of 10 locations (red dots) chosen according to Eq. 7. The locations selected are not the ones with the overall highest uncertainty, but the ones with the highest uncertainty within a neighborhood. This approach allows targeting high entropy values, while reducing information redundancy and exploring the region of interest.

The acquisition function in Eq. 7 is based on the Gaussian process upper confidence bound (GP-UCB) algorithm [34]. The GP-UBC is used in Bayesian optimization problems with an underlying Gaussian processes regression of the form *y*_*i*_ = *f* (*s*_*i*_)+*ε*_*i*_. The difference between our formulation in Eq. 7 and the original GP-UCB is that the latter uses the *mutual information* between the observations *y*_*i*_ and the process *f* [32], as opposed to the joint differential entropy of **f**_𝒜_ only. The mutual information between *y*_*i*_ and *f* is theoretically a better approach. However, the assumption of Binomial outcomes that depend on a transformation of *f*, makes this quantity harder to compute. On the other hand, the use of differential entropy showed satisfactory results in our simulation studies, as shown below.

### Experimental Simulation

To test the proposed adaptive spatial sampling approach, we conducted a series of experimental simulation studies parameterized using data from NTD surveys across multiple diseases and countries. We created different scenarios in which the task was to adaptively select new sampling locations with the goal of classifying sites as hotspots and not hotspots. In this procedure, our benchmark was the prediction performance when selecting batches of sampling sites randomly without adaptation. We defined four prevalence scenarios based on cross-sectional prevalence survey data of schistosomiasis from Cote d’Ivoire and Malawi and lymphatic filariasis from Haiti and Philippines. In each of the four countries, we used a universe of 2000 candidate survey sites identified with the *Village Finder* algorithm (see Appendix). This algorithm uses gridded population estimates of 2015 from Worldpop to identify clusters of populated places [37]. Fig 2 shows the cluster locations in each country and the simulated prevalence used as the *truth* during these experiments.

**Figure 2.**
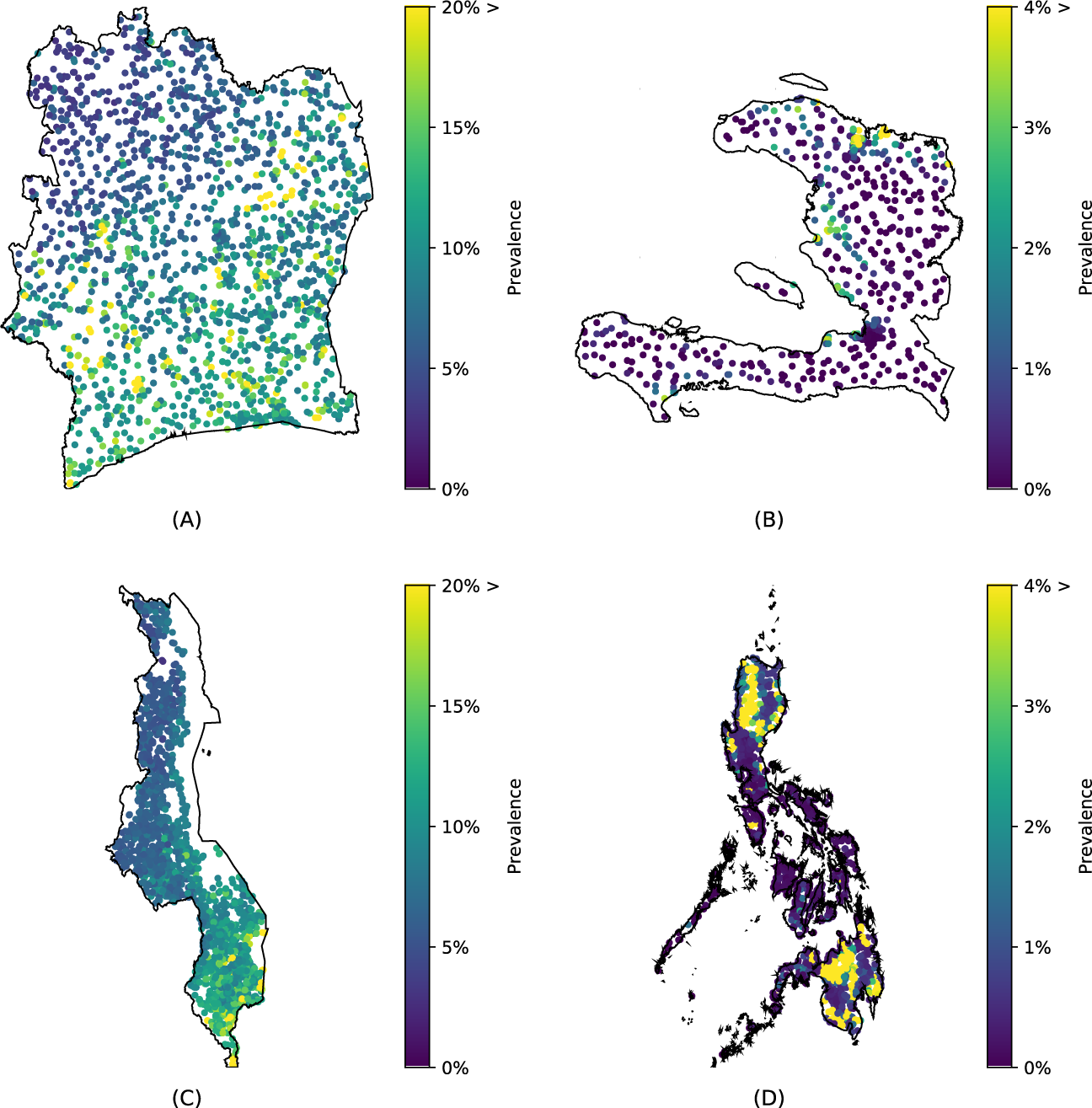
Simulated prevalence scenarios. The locations of the villages is marked by the dots, whose colors represent the hypothetical prevalence of each scenario. A: Cote d’Ivoire (schistosomiasis). B: Haiti (lymphatic filariasis). C: Malawi (schistosomiasis). D: Philippines (lymphatic filariasis).

To generate simulated prevalence estimates against which to compare different sampling approaches, a Generalized Additive Model was first fitted to the observed prevalence data from the four countries using elevation (NASA SRTM) and distance to water bodies (Digital Chart of the World) plus a spatial effect. This model was then used to predict prevalence values at every candidate cluster for each country/disease. Table 1 shows the details of the algorithm used to generate this random field of prevalence. To define hotspots, we used prevalence thresholds of 10% and 2% for schistosomiasis and lymphatic filariasis respectively, as these correspond to the thresholds used to determine whether MDA occurs or not [1, 38]. The method used to simulate these scenarios ensures that the prevalence shows spatial correlation. In addition, to ensure that we could use the standard thresholds and keep our scenarios realistic, we adjusted the overall mean of the simulated prevalence to have values around the thresholds when needed.

**Table 1.** Generation of prevalence scenarios. Based on cross-sectional surveys (line 1) and environmental data (line 2), we fitted a prevalence model (line 3). This model does not include spatial correlation explicitly, but encodes the relationship between prevalence and the variables elevation and distance to water. The linear predictors of this model were smoothed spatially (line 5) and used to generate the prevalence in all villages (lines 6 and 7). In these last two steps we explicitly included a spatial component and dropped the dependence on the environmental variables. If the mean or range of the prevalence generated was too high we scaled it to match a prevalence around the hotspot threshold of the disease they represent (line 8).

| Pseudo code for generating prevalence scenarios |  |  |
| --- | --- | --- |
| 1 | $\{y_i, n_i, s_i\}_{i=1}^{m_0} \leftarrow \text{survey data}$ | $y_i = \text{positives}, n_i = \text{total}$ |
| 2 | $\mathcal{D} = \{y_i, n_i, s_i, \mathbf{x}(s_i)\}_{i=1}^{m_0}$ | $\mathbf{x} = \text{environmental data}$ |
| 3 | fit $y_i \sim \text{Binomial}(n_i, \text{logit}^{-1}(\eta_i))$ with $\eta_i = \sum_j f_j(\mathbf{x}_{i[j]})$ | $f_j$ univariate smoother |
| 4 | $w_i = E(\eta_i \vert \mathcal{D}) \forall s_i \in \mathcal{D}$ | |
| 5 | fit $w_i = f_s(s_i) + \varepsilon_i$ with $\varepsilon_i \sim \mathcal{N}(0, \sigma^2)$ | $f_s$ spatial smoother |
| 6 | predict $w_i^* \forall s_i \in \mathcal{S}$ | $\mathcal{S} = \text{villages in the country}$ |
| 7 | $\theta_i = \text{logit}^{-1}(w_i^*)$ | $\theta_i = \text{simulated prevalence}$ |
| 8 | $\theta_i \leftarrow \text{check values and adjust if needed}$ | |

In order to have consistency in our results, we repeated our experiments 50 times per country. In each replicate, we randomly selected 100 locations from the universe of clusters and used them as the locations of the initial set 𝒟_0_. We ran three versions of the experiments, by sequentially selecting batches of size 1, 10 and 50, until we had incorporated 100 new samples. Given a set of initial sampling locations and batch size, we sampled additional locations either completely at random or adaptively following Eq. 7. At each step we fitted the model described in Eq. 1 to 3. As environmental variables we used: annual mean temperature, temperature seasonality, annual precipitation and precipitation seasonality [39], elevation (SRTM) and distance to inland water resampled to the same 1km resolution. After fitting the spatial model on each iteration, we computed four out-of-sample validation statistics to measure performance (see Appendix): accuracy, positive predictive value (PPV), sensitivity and mean squared error (MSE). To compute the validation statistics we fitted the model in Eq. 1 to all the available data at each iteration (i.e. 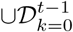 at step *t*) and made predictions on the villages that had not been visited yet (i.e.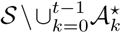 at step *t*). MSE was computed comparing the predicted prevalence vs the simulated prevalence (see Table 1). To compute accuracy, positive predictive value and sensitivity we first classified the villages as hotspots when *p*(*θ*_*i*_ *> ϑ*|𝒟_0_) *>* 0.5 and compared this classification vs the actual class according to the simulated prevalence. Table 2 shows the algorithm followed to carry on our experiments.

**Table 2.** Experimental procedure. We repeated each experiment a hundred times (line 1), for batches of size 1, 10 and 50 (line 2). We started with an initial random sample of 100 locations (line 3) for both random and adaptive methods (line 4). We incorporated subsequent samples until 100 additional sampling locations were added (line 5). For the locations selected to be sampled we simulated the observed positive cases according to a Binomial distribution with prevalence *θ* (line 7) and incorporated the environmental data (line 8). We then used the accumulated data to find the probability of exceeding the threshold *ϑ* (line 9). Finally we defined a new batch of locations according to a random mechanism (line 11) and to the adaptive sampling method proposed (line 12).

| Pseudo code for experiments |  |  |
| --- | --- | --- |
| 1 | for rep in 1 → 100: | |
| 2 | for m in {1, 10, 50}: | $m = \text{batch size}$ |
| 3 | $\mathcal{A}_0 \leftarrow \text{random selection: } \mathcal{A}_0 \subset \mathcal{S} \text{ with } \ \mathcal{A}_0\ = 100$ | $\mathcal{S} = \text{villages in the country}$ |
| 4 | $\mathcal{A}_0^R = \mathcal{A}_0^A = \mathcal{A}_0$ | $R = \text{random}, A = \text{adaptive}$ |
| 5 | $\text{steps} = 100/m + 1$ | $\text{total number of iterations}$ |
| 6 | for t in 1 → steps: | |
| 7 | $y_i^* \sim \text{Binomial}(100, \theta(\mathcal{A}_{t-1}^*))^\dagger$ | $* = \{R, A\}$ |
| 8 | $\mathcal{D}_{t-1}^* = \{\mathcal{A}_{t-1}^*, y^*, \mathbf{x}(\mathcal{A}_{t-1}^*)\}$ | $\mathbf{x} = \text{environmental data}$ |
| 9 | find $p(\theta > \vartheta \vert \cup_{k=0}^{t-1} \mathcal{D}_k^*)$ | |
| 10 | compute validation statistics on $\mathcal{S} \setminus \cup_{k=0}^{t-1} \mathcal{A}_k^*$ | |
| 11 | $\mathcal{A}_t^R \leftarrow \text{random selection}$ | |
| 12 | $\mathcal{A}_t^A \leftarrow \text{acquisition function Eq. 7}$ | |

$^\dagger y_i^R = y_i^A$ for step $t = 0$ .

Random forest and geostatistical models were fit using the R packages ranger [40] and spaMM [41] respectively. All the simulated datasets and code developed as part of this study, including that used to conduct the simulation experiments, is available at https://github.com/disarm-platform/adaptive_sampling_simulation_r_functions.

We are also in the process of developing a user-friendly web application to allow both the hotspot mapping and adaptive sampling algorithms to be run without code.

## Results

We compared the performance of two approaches for selecting survey sites: random sampling (RS), where sites are chosen randomly; and adaptive sampling (AS), that follows the acquisition function of Eq. 7. The underlying statistical model is the same in both cases (see Eq. 1 - 3). The initial dataset 𝒟_0_ is also the same in both cases (see Table 2 lines 7 and 8). Hence, the variations in the performance with respect to the predictions based on 𝒟_0_ depend only on the mechanism of selecting the new survey locations 𝒜_1_, 𝒜_2_,…. Adding measurements at new locations improved out-of-sample sites classification under both sampling approaches. However, across the four scenarios tested we observed that adaptive sampling was consistently superior to random sampling in terms of accuracy, positive predictive value and sensitivity. This confirms that under adaptive sampling each new batch of locations leads to a better classification of the unmeasured sites. Figure 3 shows the accuracy computed at each step in the four country scenarios using a batch of size 1. Note that when selecting a batch of size 1, the adaptive design does not take into account the exploration component. In this case the new location suggested is the one that maximizes entropy. Figure 4 shows a summary of the validation statistics after adding 100 new samples, using different batch sizes (1, 10 and 50), across the four scenarios. The results show that adaptive sampling produces superior accuracy, sensitivity and PPV across every scenario, metric and batch size except in the Philippines where an adaptive approach with a batch size of 50 produced inferior PPV. Better performance across all metrics translates into a smaller number of false positives and a improved identification of hotspots in locations that have not been visited yet. In contrast to the validation statistics discussed above, MSE (bottom row) is lower across all scenarios when random sampling was employed.

**Figure 3.**
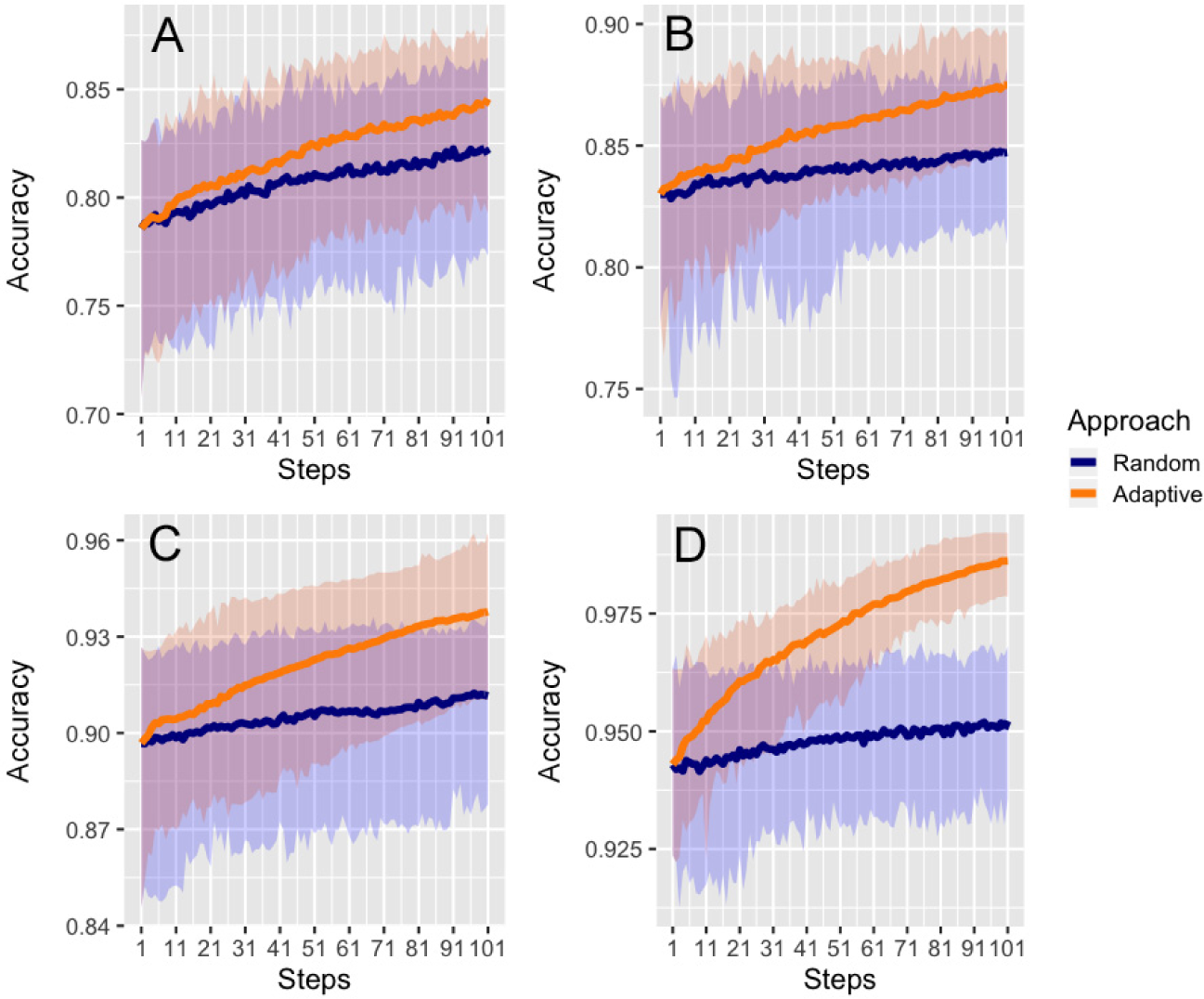
Out of sample accuracy (batch size = 1). The solid line represents the average value across 50 repetitions. The shaded area represents the 2.5% and 97.5% quantiles of the values observed across all 50 repetitions at each step. A: Cote d’Ivoire (*ϑ* = 10%). B: Malawi (*ϑ* = 10%). C: Haiti (*ϑ* = 2%). D: Philippines (*ϑ* = 2%).

**Figure 4.**
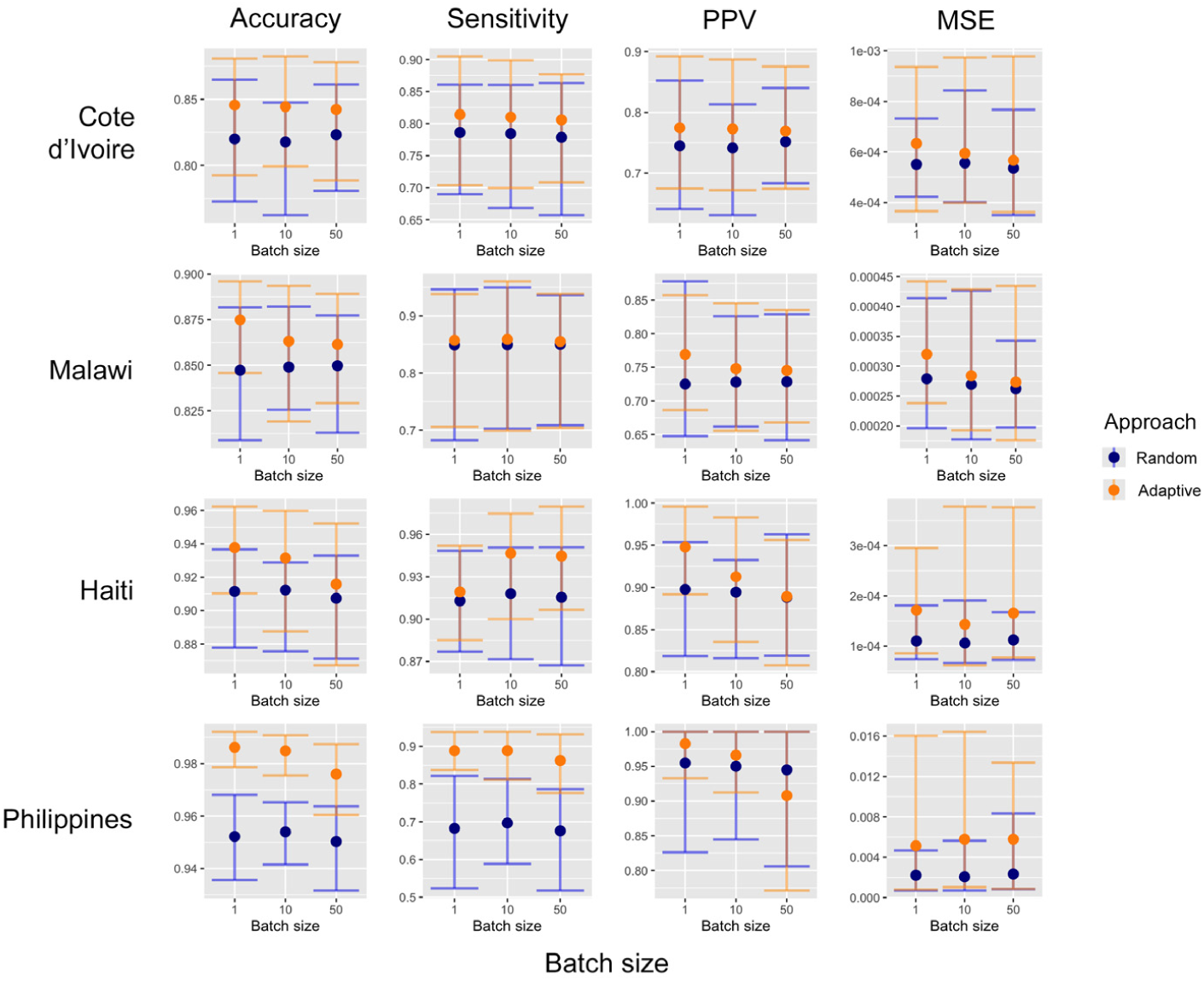
Summary of validation statistics. Metrics computed after adding 100 new samples in batches of 1, 10 and 50 sites. Dots represent the mean and whiskers represent the the 2.5% and 97.5% quantiles of values observed across all 50 repetitions. The thresholds used to define a hotspot are: *ϑ* = 10% in Cote d’Ivoire and Malawi and *ϑ* = 2% in Haiti and Philippines.

At larger batch sizes there were smaller differences between random and adaptive sampling in terms of accuracy, PPV and sensitivity (Figure 4). There are two ways of interpreting this result. One interpretation is that when the batch is large enough, random sampling provides a good coverage of the sampling universe negating the need for a trade-off between exploitation and exploration. The more locations in the batch the more redundant the information they provide, regardless of how they are chosen. A second interpretation is that the adaptive sampling design is more efficient and therefore requires smaller sample sizes to achieve the same results of a larger random sample. Table 3 illustrates this and shows the number of sample points needed when using adaptive sampling to achieve the same accuracy of random sampling with a sample size of 100 locations. For batches of size 50, in Malawi and Philippines adaptive sampling produced at least the same level of accuracy with just half the number of additional points. This difference becomes larger for smaller batch sizes (1 or 10). For batch sizes of 10, adaptive sampling required 20-50% of the sample size to achieve the level of accuracy achieved with 100 additional randomly selected sites and for batch sizes of 1, only 10-43% was required. With such sample sizes the adaptive sampling also achieved similar levels of sensitivity and PPV. Random sampling produced lower MSE across all scenarios.

**Table 3.** For random design RS with sample size of 100, we show the sample size needed to achieve a similar accuracy using an adaptive design AS. Additional validation statistics: PPV, sensitivity and MSE are also shown. Along the rows, results are shown per country and batch size ‖*A*_*i*_‖.

| Country | $\ \mathcal{A}_i\ $ | Num. obsv. | | Accuracy (%) | | PPV (%) | | Sensitivity (%) | | MSE ($\times 10^{-4}$) | |
| --- | --- | --- | --- | --- | --- | --- | --- | --- | --- | --- | --- |
|  |  | RS | AS | RS | AS | RS | AS | RS | AS | RS | AS |
| Cote d'Ivoire | 1 | 100 | 43 | 82.0 | 82.0 | 78.6 | 77.7 | 74.5 | 74.7 | 5.5 | 6.8 |
|  | 10 | 100 | 50 | 81.7 | 82.4 | 78.4 | 77.9 | 74.1 | 75.3 | 5.6 | 6.4 |
|  | 50 | 100 | 100 | 82.3 | 84.2 | 77.8 | 80.6 | 75.2 | 76.9 | 5.4 | 5.7 |
| Malawi | 1 | 100 | 26 | 84.7 | 84.9 | 84.9 | 82.7 | 72.5 | 73.4 | 2.8 | 3.4 |
|  | 10 | 100 | 40 | 84.9 | 84.9 | 85.0 | 84.3 | 72.8 | 72.9 | 2.7 | 3.1 |
|  | 50 | 100 | 50 | 85.0 | 85.0 | 85.0 | 83.9 | 72.9 | 73.1 | 2.6 | 3.0 |
| Haiti | 1 | 100 | 26 | 91.1 | 91.3 | 91.3 | 91.1 | 89.8 | 90.4 | 1.1 | 1.5 |
|  | 10 | 100 | 40 | 91.2 | 91.5 | 91.8 | 92.6 | 89.4 | 89.6 | 1.1 | 1.8 |
|  | 50 | 100 | 100 | 90.7 | 91.6 | 91.5 | 94.5 | 88.8 | 88.9 | 1.1 | 1.7 |
| Philippines | 1 | 100 | 10 | 95.2 | 95.2 | 68.2 | 67.7 | 95.5 | 94.4 | 2.2 | 4.7 |
|  | 10 | 100 | 20 | 95.4 | 95.8 | 69.7 | 71.7 | 95.0 | 94.1 | 2.0 | 4.6 |
|  | 50 | 100 | 50 | 95.0 | 95.5 | 67.6 | 79.3 | 94.5 | 83.5 | 2.3 | 5.7 |

## Discussion

The identification of disease hotspots is an increasingly important public health problem. This is particularly true in disease elimination settings, where transmission is rare and typically focal. Numerous examples illustrate the use of geospatial modeling to predict hotspots, but very little attention has been given to the optimal survey design for such modeling efforts. Here, using simulation studies based on schistosomiasis and lymphatic filariasis survey data, we described a novel, spatially adaptive approach and demonstrate the superiority of this approach at identifying hotspots compared with the standard approach to surveys based on purely random sampling.

Results showed that across all batch sizes investigated, adaptive approaches produced higher levels of accuracy, sensitivity, and PPV compared with random sampling. Yet, the superiority of an adaptive approach declined with larger batch sizes. With a batch size of 1, the adaptive approach has an opportunity to identify the optimal next location to survey in the presence of all available data. In contrast, with larger batch sizes, the impact on predictions of each adaptively sampled location is not known until all locations in the batch are sampled and the model updates. The use of an adaptive approach only produced marginal gains in accuracy (1-4%) after adding 100 sites to the initial sample, but this could represent hundreds of locations when applied at a country scale. Perhaps more importantly, however, adaptive sampling was more efficient in terms of achieving a given level of accuracy with a far smaller sample size. As outlined in Table 3, across almost all settings, adaptive sampling was able to achieve the same level of accuracy and sensitivity to that achieved by adding 100 locations randomly with between 10-50% the sample size.

The only exception to this was using batch sizes of 50 in Cote d’Ivoire and Haiti where 100 adaptively selected sites were required to achieve at least the same performance as an additional 100 randomly selected sites. These results demonstrate that an adaptive spatial sampling approach has the potential to substantially reduce the resources required to ensure hotspot locations receive treatment, while maintaining similar rates of false positives. In control and elimination settings, an operationalized adaptive spatial sampling approach for several years could render non-negligible improvements in cost-effectiveness. Further simulation studies could be used to help determine the magnitude of such benefits in cost-effectiveness.

It should also be pointed out that in all settings the mean squared error estimates were higher for adaptive approach (Figure 4, Table 3). This illustrates the fact that optimizing a design for one goal, here hotspot classification accuracy, leads to compromising other goals (e.g. precision in the prevalence estimates). Where the goal is to produce the most precise prevalence estimates at any given location, using adaptive approaches based on prediction variance as opposed to entropy would be more appropriate [25].

While this approach was demonstrated for two diseases only, it could be used to support the identification of hotspots of any binomial outcome. This includes prevalence of infection of other infectious and non-infectious diseases, particularly those that display strong spatial correlation. Vector-borne diseases, such as malaria, onchocerciasis and loiasis would certainly fall into this category given the association between disease transmission and ecological and environmental conditions. While it is likely that such spatial correlation will be masked following several years of intervention, evidence suggests that residual hotspots still occur [2, 42]. In addition to identifying hotspots of infection, this approach also has potential utility for identifying *cold spots* in intervention coverage, such as pockets of undervaccination [43]. While this would likely require use of different covariates related to intervention access, such as distance to roads, population density and poverty, the statistical problem is analogous.

While we used a combination of random forest and model-based geostatistics to produce posterior prevalence estimates, the general adaptive sampling scheme we have proposed would work for any suitable modeling approach that produces posterior estimates with which to estimate exceedance probabilities. Combining random forests with other base learners such as generalized additive models and support vector machines may lead to improvements over using random forest alone. Similarly, an underlying binomial model is not essential to the methodology described here. What is important is the spatial correlation component in which the exploration rule is based. For example, this methodology could work in a Poisson setting, for some definition of hotspot based on a threshold incidence or numbers of cases.

Another possible extension of this methodology is applying it to cases where the classification of interest is not binary. For example, for schistosomiasis, MDA is recommended once per year in areas where prevalence is *>* 10% and *<* 50% and twice per year in areas where prevalence is *>* 50% [1]. As estimation of entropy is not restricted to binary classification problems, adapting the approach to such a setting is straightforward assuming it is possible to produce probabilistic classifications from the underlying model.

This study had a number of limitations. Firstly, the adaptive sampling approach described requires a georeferenced set of candidate sampling locations. Complete georeferenced lists of settlements are, however, often not available. In the absence of such data, there are several options available. Georeferenced locations could be extracted and compiled from open sources, such as openstreetmap, geonames and openAFRICA. Alternatively, village locations can be derived from gridded population data using the approach described here (see Appendix) or using alternative approaches as suggested by Thompson et al (2017) [44].

A second limitation is that we did not consider the temporal aspect of adaptive surveys. In reality, there may be a time lag between the date at which survey data are available and when adaptive surveys take place. Similarly, prior survey data may have been collected over multiple time periods. To address this issue it would be possible to extend the spatial model used, to a spatio-temporal model. Hotspot probabilities could then be forecast from the historic data to the time point at which adaptive surveys are to take place. Additionally, there may be value in using temporally dynamic covariates as opposed to static, long-term averages as used here.

A third limitation was that we defined a site as hotspot if there was at least a 50% chance that prevalence exceeded the relevant threshold. In some cases, programs may *a priori* wish to define hotspots more conservatively by classifying sites as hotspots with smaller probabilities (e.g. *>* 10% chance a site is a hotspot). While the methodology would not change, such an approach would have a large impact on the performance of the classifications, increasing sensitivity, but decreasing positive predictive value. In such cases, it may also be useful to modify the acquisition function.

A fourth limitation of this study is that we used a single acquisition function. In the acquisition function we used, the exploration component has an increasing concave weight as more locations are added to the new batch. This assumption, or the specific shape of this weight, could be substituted for an alternative. Also, the utility function, defined here as entropy, could be modified depending on the goal pursued. For example, a program interested in targeting sampling efforts at hotspots, instead of achieving a better binary classification, could use the probability of a location being a hotspot as the utility function. Such an approach would be suitable for situations where testing is required before an intervention/treatment is administered. This approach may also be useful for surveys whose goal is to determine freedom from infection [45, 46].

A fifth limitation stems from the simulated nature of the experiments. The strength of a simulated approach is that multiple experiments can be conducted without the need for expensive field validation studies. On the basis of these results, a valuable next step would be to conduct field studies comparing random to adaptive designs. Such studies would also allow an exploration of some of the more logistical elements and constraints and using an adaptive approach.

This study has demonstrated the value in adopting an adaptive approach to surveys designed to identify disease hotspots. Results show that a spatially adaptive sampling approach produced consistently superior accuracy in hotspot classification over a random sampling approach, and could dramatically lower the resources requirements to conduct surveys whose goal is to detect disease hotspots.

## Data Availability

All the simulated datasets and code developed as part of this study, including that used to conduct the simulation experiments, is available on a public GitHub repository.

https://github.com/disarm-platform/adaptive_sampling_simulation_r_functions

## Author’s contributions

HJWS, RAP and BA conceived the study. RAP, FR and HJWS conducted all statistical analyses. RAP and HJWS lead the writing of the manuscript. JL, LH, AM and LJ oversaw collection of the survey data and AB and ML provided input on the statistical approach and on the manuscript.

## Acknowledgements

This work received financial support from the Coalition for Operational Research on Neglected Tropical Diseases, which is funded at The Task Force for Global Health primarily by the Bill & Melinda Gates Foundation, by the United States Agency for International Development through its Neglected Tropical Diseases Program, and with UK aid from the British people.

## Appendix Covariance Structure

In Eq. 1 - 3, it is the function *f* (**s**_*i*_) the one that encodes the spatial structure. Here we model such spatial structure as an *Matérn* covariance, given by

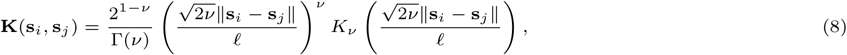

where *ν* controls the smoothness of the process, 𝓁 is a lengthscale parameter and *K*_*ν*_ is a modified Bessel function.

### Village Finder

The Village Finder algorithm is accessible via a Shiny app that suggests GPS coordinates of populated sites based on 1km resolution Worldpop gridded population data. A populated site is an area that meets certain size and population criteria and can represent a village, a neighborhood of a crowded city or a large but sparsely populated rural area. The user specifies the following 3 parameters to define the type of population sites queried:

- maximum area size, above which a region cannot be considered as a unique location;
- upper population threshold, above which a location should be counted as a unique location;
- lower population threshold, below which a region smaller than the maximum area size should not be counted as a populated location.

The algorithm works iteratively. First, any 1km grid cells of the Worldpop raster that adhere to the three parameters are identified and the centroids are kept. The gridded population data, minus those grid cells identified in the first round, are then aggregated by a factor of 2 and any aggregated areas that adhere to the parameters are identified. The centroid of the most populated cell in the aggregated area is then assigned as the village location for that aggregated area. The process continues until all aggregated areas have an assigned centroid or until all thresholds are met.

This app and the code behind are available from:

- https://disarm.shinyapps.io/ui-village-finder;
- https://github.com/disarm-platform/fn-village-finder.

#### Validation Statistics

To measure the performance of the classification model we used four different metrics. To define them, we first need to define the following terms:

- True positives (*tp*): cases where the actual category and the predicted category are both positive (e.g. a site classified as a hotspot actually has a prevalence above the threshold of interest).
- True negatives (*tn*): cases where the actual category and the predicted category are both negative (e.g. a site classified as not being a hotspot actually has a prevalence below the threshold of interest).
- False positives (*fp*): cases where the actual category is negative, but the predicted class is positive (e.g. the site is classified as a hotspot, but the actual prevalence is below the threshold of interest).
- False negatives (*fn*): cases where the actual category is positive, but the predicted class is negative (e.g. the site is classified as not being a hotspot, but the actual prevalence is above the threshold of interest).

#### Accuracy

The proportion of sites correctly classified.

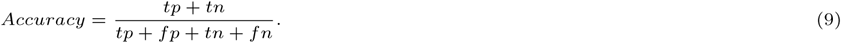

#### Positive predicted value

The proportion of sites correctly classified as hotspots to all sites classified (correctly or incorrectly) as hotspots.

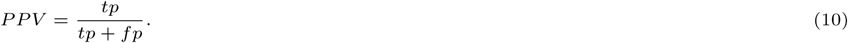

#### Sensitivity

The proportion of sites correctly classified as hotspots to all hotspots in the dataset.

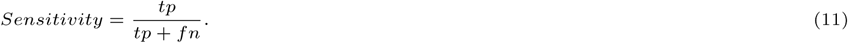

#### Mean squared errors

The average of the squared differences between the target value (predicted prevalence) and the observed value (actual prevalence).

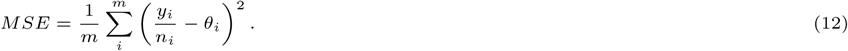

